# Colorectal cancer incidence and mortality trends by age at onset in South Africa from 2002 to 2023

**DOI:** 10.64898/2026.11.16.26368230

**Authors:** Wenlong Carl Chen, Jean-Tristan Brandenburg, Sizeka A. Mashele, Natasha Abraham, Babongile Ndlovu, Matshediso I. Mohlala, Tinashe Tombe, Judith R.M. Mwansa-Kambafwile, Carole Metekoua Temdemnou, Zanele Mtsweni, Nchimane Mahladisa, Vusi Nzimakwe, Mosidi Nhlapo, Paul Ruff, Philip C. Haycock, the MOSAIC collaboration, Mazvita Muchengeti

## Abstract

**Background:** Colorectal cancer is the third most common cancer in men and women, and the fifth most common cancer overall, in South Africa. A previous report by the South African National Cancer Registry (SANCR) established that colorectal cancer incidence is rising in all South African population groups but rising fastest in Black South Africans. However, whether incidence increases by age at onset, as observed in many upper-income countries worldwide, remains unknown. In the present study, we assessed changes in colorectal cancer incidence (2002–2023) and mortality (2002–2019) in South Africa by sex, population group and age at onset.

**Methods:** We calculated incidence from SANCR case counts and mortality from Stats SA death records, using Stats SA mid-year population estimates, standardised to the Segi world standard population. We defined age at onset as early (<50 years) or late (≥50 years). We described trends using joinpoint regression and annual percentage changes (APCs) over 2002–2023 and 2011– 2023, the period after private-laboratory reporting became mandatory.

**Results:** A total of 70,467 histologically confirmed primary colorectal cancers were registered with the SANCR over 2002–2023; 53.7% were in men. The overall age-standardised incidence rate (ASIR) was 6.61 per 100,000 (95% CI 6.32–6.89) in 2002 and 8.23 (7.99–8.46) in 2023. Overall incidence fell from 2002 to 2007 (not significant), rose from 2007 to 2015 and showed no significant change from 2015 to 2023. The first two segments coincide with the withdrawal of some private laboratories from reporting in 2005–2007 and with mandatory reporting from 2011. From 2011 to 2023, incidence fell in White South Africans (APC −1.0%, 95% CI −1.9 to −0.04), rose in Black African South Africans (+3.1%, 2.0 to 4.2) and did not change significantly in Coloured (−1.0%, −2.3 to 0.3) or Indian/Asian (+0.3%, −2.2 to 2.7) South Africans. Early-onset incidence rose by 1.6% per year (95% CI 1.1 to 2.1) over 2002–2023, similar to late-onset incidence (1.8%, 1.0 to 2.5). After 2011, neither changed significantly (early-onset +0.3%, −0.3 to 0.9; late-onset 0.0%, −1.0 to 1.0). Over 2002–2023, the early-onset APC was highest in White South Africans (+3.9%, 2.9 to 4.9), followed by Black African (+1.7%, 1.1 to 2.4), Coloured (+1.6%, 1.0 to 2.3) and Indian/Asian (+1.5%, 0.4 to 2.7) South Africans. The late-onset APC was +4.1% (3.4 to 4.9) in Black African, +1.3% (0.1 to 2.5) in White, +1.3% (0.1 to 2.5) in Indian/Asian and +0.7% (−0.3 to 1.6) in Coloured South Africans. After 2011, early-onset incidence rose in White (+1.2%, 0.3 to 2.2), Coloured (+1.7%, 0.4 to 3.1) and Black African (+1.1%, 0.4 to 1.8) South Africans. Late-onset incidence fell in White South Africans (−1.5%, −2.6 to −0.4) and rose in Black African South Africans (+3.7%, 2.4 to 5.0). Early-onset mortality rose by 1.2% per year (0.5 to 2.0) over 2002–2019, and late-onset mortality by 0.9% (0.3 to 1.4).

**Conclusion:** Early-onset colorectal cancer incidence in South Africa rose over 2002–2023, and much of the rise coincided with mandatory private-laboratory reporting from 2011. Since 2011, early-onset incidence has risen modestly in White, Coloured and Black African South Africans, while late-onset incidence has fallen in White South Africans. In Black African South Africans, late-onset incidence and mortality continue to rise faster than early-onset.

## 1. Background

Colorectal cancer is the third most common cancer in men and women, and the fifth most common cancer overall, in South Africa^1^. Globally, colorectal cancer is the third most common cancer and is the second leading cause of cancer mortality^2^. Across most examined countries, incidence rates are rising faster for early (<50 yrs) compared to late (≥50 yrs) onset colorectal cancer^3^. In a previous report by the South African National Cancer Registry (SANCR), colorectal cancer incidence was highest in White South Africans but was rising fastest in Black Africans^4^. However, whether the rising incidence varies by age-at-onset was not assessed. Clarifying changes in incidence by age-at-onset could have important policy implications, given that early-onset cases account for a relatively high proportion of incident colorectal cancers in Black South Africans (35%^5^ versus 9.6% globally^6^) and median age-at-diagnosis is 10 years earlier than in White South Africans (54.5 versus 64.5)^4^. Although these age-at-onset statistics probably reflect differences in population age structure and life expectancy, mismatch repair deficiency is more common in tumours from individuals of African ancestry and early-onset cases, pointing to potential biological explanations^6,7^.

Established in 1986, the SANCR is a pathology-based monitoring system that collected data on cancer from private and non-private laboratories on a voluntary basis up until 2011. Interpretation of statistics from the SANCR requires careful consideration of a number of reporting disruptions that have occurred over the past two decades. For example, owing to the voluntary nature of data sharing, some private sector laboratories stopped sharing data with the SANCR from 2005 to 2007^8^. In 2011, legislative changes made data reporting to the SANCR compulsory for all pathology laboratories^9^. In 2015, the SANCR changed its case-finding procedures (SANCR, unpublished). The COVID-19 pandemic in 2020 caused further disruptions to case reporting. SANCR data must also be interpreted in the context of substantial inequality in health care access amongst its population groups. South Africa has a dual health system: 16% of the population have private healthcare insurance, while 84% obtain health care from the government sector^8^. Economically, South Africa is one of the most unequal societies in the world, with a Gini coefficient of 0.64^10^. Due to its political history, inequality is stratified by population groups, defined by Statistics South Africa (StatsSA) as Black African (81.1%), Coloured (8.8%), White (7.5%) and Indian/Asian (2.6%) in 2023^11^. 72% of the White population has private health insurance, followed by Indian/Asian (41%), Coloured (20%) and Black African (10%)^12^. Although a minority of Black Africans have private health insurance, Black Africans account for 51% of members of private health care schemes^12^.

Previous studies by the SANCR have assessed incidence and mortality trends for colorectal cancer by sex, population group and anatomical site^4^ but not by age-at-onset. The main aim of the present study was to update these previous reports by focusing on rates of change in early-onset and late-onset colorectal cancer incidence and mortality among population groups in South Africa. The idea to conduct this updated analysis emerged from the Wits-Bristol colorectal cancer workshop, held at the University of the Witwatersrand in Johannesburg (30^th^ September to 1^st^ October 2025). The workshop brought together clinicians, patient representatives and scientists from South Africa, Botswana, the UK and France to discuss colorectal cancer research priorities in southern Africa. The workshop established the MOSAIC collaboration, which aims to uncover novel MOlecular, Somatic And Infectious drivers of colorectal cancer.

## 2. Methods

### 2.1 Study Design and Data Source

This was a retrospective, population-based study of incidence and mortality trends, using data from the SANCR and Stats SA. The SANCR is a pathology-based cancer registry: it compiles records exclusively from histopathological reports submitted by public and private diagnostic laboratories throughout South Africa. It does not capture cases identified solely by clinical or imaging criteria. All histologically confirmed primary colorectal cancers with ICD-O-3 topography codes C18 (colon), C19 (rectosigmoid junction), and C20 (rectum), and behaviour code /3 (malignant primary), diagnosed between 2002 and 2023, were eligible for the incidence analysis.

### 2.2 Case Ascertainment and Inclusion Criteria

For the incidence analysis, records were included if they met all of the following criteria: (i) ICD-O-3 topography C18.0–C20.9 with behaviour code /3; (ii) year of diagnosis 2002–2023; (iii) known age at diagnosis; and (iv) known sex (male or female). Records with unrecorded population group (2,318 cases, 3.3%) were included in overall, sex-specific and onset-specific rates, because the population denominator includes all population groups. They were excluded from population-group-specific rates.

Population group classification follows the South African national statistical convention and is used solely as a demographic stratifier consistent with published SANCR data and Stats SA denominators.

### 2.3 Study Period and Population Denominators

The primary incidence analysis period was 2002 to 2023, encompassing all years for which final SANCR registration data and confirmed Stats SA population denominators were available. Denominators for 2002–2022 were obtained from the published Stats SA mid-year population estimate series; 2023 denominators were extracted from the official Stats SA 2023 report, confirmed as final before inclusion. Denominators were structured as 128 cells per year (16 five-year age bands from 0–4 to 75+, four population groups, and two sexes).

The expansion of private-sector laboratory reporting to the SANCR from approximately 2011 onwards led to a structural step increase in registered case counts, reflecting improved case ascertainment rather than a true change in incidence. This limitation is acknowledged throughout and is addressed in sensitivity analyses SA1 and SA3.

### 2.4 Age-Standardised Incidence Rates

Age-standardised incidence rates (ASIRs) were computed using the Segi-Doll world standard population (total = 100,000 person-years). A two-step computation was applied to avoid aggregation bias: cases and person-years were first summed within each five-year age band across sub-strata; age-specific rates were then computed and weighted by the corresponding Segi weight. Formally:

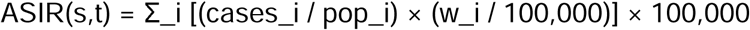

where i indexes five-year age bands, and w_i denotes the Segi weight. The variance of each rate was estimated assuming Poisson-distributed counts, and 95% confidence intervals (CIs) were calculated as ASIR ± 1.96 × SE. Truncated ASIRs were computed for age-at-onset analyses: ages 0–49 for early-onset and 50 and older for late-onset. The Segi weights for these age ranges sum to 80,000 and 20,000, respectively, and were rescaled to sum to 100,000.

### 2.5 Age-at-Onset Classification

Colorectal cancer was classified as early-onset if the age at diagnosis was below 50 years, and as late-onset if 50 years or older, consistent with the threshold used in most international studies of early-onset colorectal cancer^13,14^.

### 2.6 Joinpoint Regression

Temporal trends in overall, sex-stratified, onset-stratified, and population group–specific ASIRs were modelled using the NCI Joinpoint Regression Program v6.0.1 (National Cancer Institute, USA): log-linear model, maximum four joinpoints, 4,499 Monte Carlo permutations, with heteroscedastic errors based on the standard error of each rate. The annual percentage change (APC) and its 95% confidence interval are reported for each segment, with confidence intervals from the empirical quantile method; * denotes an APC whose 95% CI excludes zero. Because the 2011 reporting mandate falls inside the study period, we also report period APCs for 2002–2023 and 2011–2023 (mortality 2002–2019 and 2011–2019). Each was estimated by weighted least-squares regression of the log rate on calendar year, with weights equal to the inverse variance of the log rate and the variance scaled by the residual dispersion.

### 2.7 Regression Modelling

Poisson regression was fitted to counts by calendar year, five-year age band (15 years and older) and sex, with the log of the population as offset, age band and sex as covariates, and separate year slopes for early- and late-onset disease. Variance was estimated with robust (sandwich) standard errors because of overdispersion (Pearson dispersion 1.3 to 4.3). The difference between the onset-specific slopes was tested with a Wald test. Models were fitted for all population groups combined and for each group separately.

### 2.8 Sensitivity Analyses

Three pre-specified sensitivity analyses were conducted:

- **SA1: Restricted 2011–2023,** re-analysis confined to the comprehensive private-sector reporting era.
- **SA2: Exclusion of 2020,** re-analysis excluding the COVID-19 pandemic year, during which registration disruption may have caused artificial underreporting.
- **SA3: a level-shift indicator at 2011** (and, if confirmed, at the case-finding change) added to the primary model.

### 2.9 Mortality, mortality-to-incidence ratio and provincial analysis

Deaths with an underlying cause of C18–C20 were obtained from Stats SA death records for 2002– 2022; stillbirth records were excluded. Deaths with an unrecorded population group (6,742; 14.3%) were included in overall, sex-specific and onset-specific rates and excluded from population-group-specific rates. Because deaths fell sharply in 2020 and had not returned to the 2019 level by 2022, the primary mortality window was 2002–2019. Age-standardised mortality rates (ASMR) used the same standard and truncated weights as incidence. The mortality-to-incidence ratio (MIR) was calculated as ASMR / ASIR for each year. Provincial ASMR was calculated for 2006–2019 using province of residence, with province of death where residence was missing; provincial populations were derived from Stats SA P0302 provincial shares raked to the national denominator. Provincial rates are age-standardised but not adjusted for population group, because no provincial population estimates by population group are published. Provincial incidence could not be calculated because the SANCR records the reporting laboratory rather than the patient’s residence.

### 2.10 Ethical Considerations

This study used de-identified routinely collected registry data. Individual participant consent was not required. The National Cancer Registry of South Africa conducted this study under the ethi al waiver (W-CBP-230118-01) granted by the University of the Witwatersrand’s Human Research Ethics Committee. This waiver allows the National Cancer Registry to fulfil its legislative mandate for cancer surveillance in South Africa and to support related research. The study was conducted in accordance with the Declaration of Helsinki.

## 3. Results

### 3.1 Overall Incidence Trends (2002–2023)

A total of 70,467 histologically confirmed primary colorectal cancer cases were registered with the SANCR over the 22-year primary analysis period (2002–2023). Annual case counts increased from 2,106 in 2002 to 4,665 in 2023, with the largest annual increase from 2010 to 2011 (2,385 to 3,042 cases), coinciding with mandatory private-sector laboratory reporting (Supplementary Figure S1). The overall age-standardised incidence rate (ASIR) fluctuated between 5.59 per 100,000 (2007; 95% CI 5.35–5.84) and 8.97 (2017; 8.69–9.24). The 2002 ASIR was 6.61 (6.32–6.89), and the 2023 ASIR was 8.23 (7.99–8.46).

Joinpoint regression identified two change points in the combined-sex trend, in 2007 (95% CI 2004– 2012) and 2015 (2012–2018) (Figure 1; Table 1). Incidence did not change significantly from 2002 to 2007 (APC −2.63%, 95% CI −12.54 to 1.81), rose from 2007 to 2015 (+5.40%, 3.35 to 13.45) and did not change significantly from 2015 to 2023 (−1.24%, −3.58 to 0.51). The 2007–2015 rise in ASIR coincides temporally with the expansion of private-sector reporting and should be interpreted in the context of improved ascertainment.

**Figure 1.**
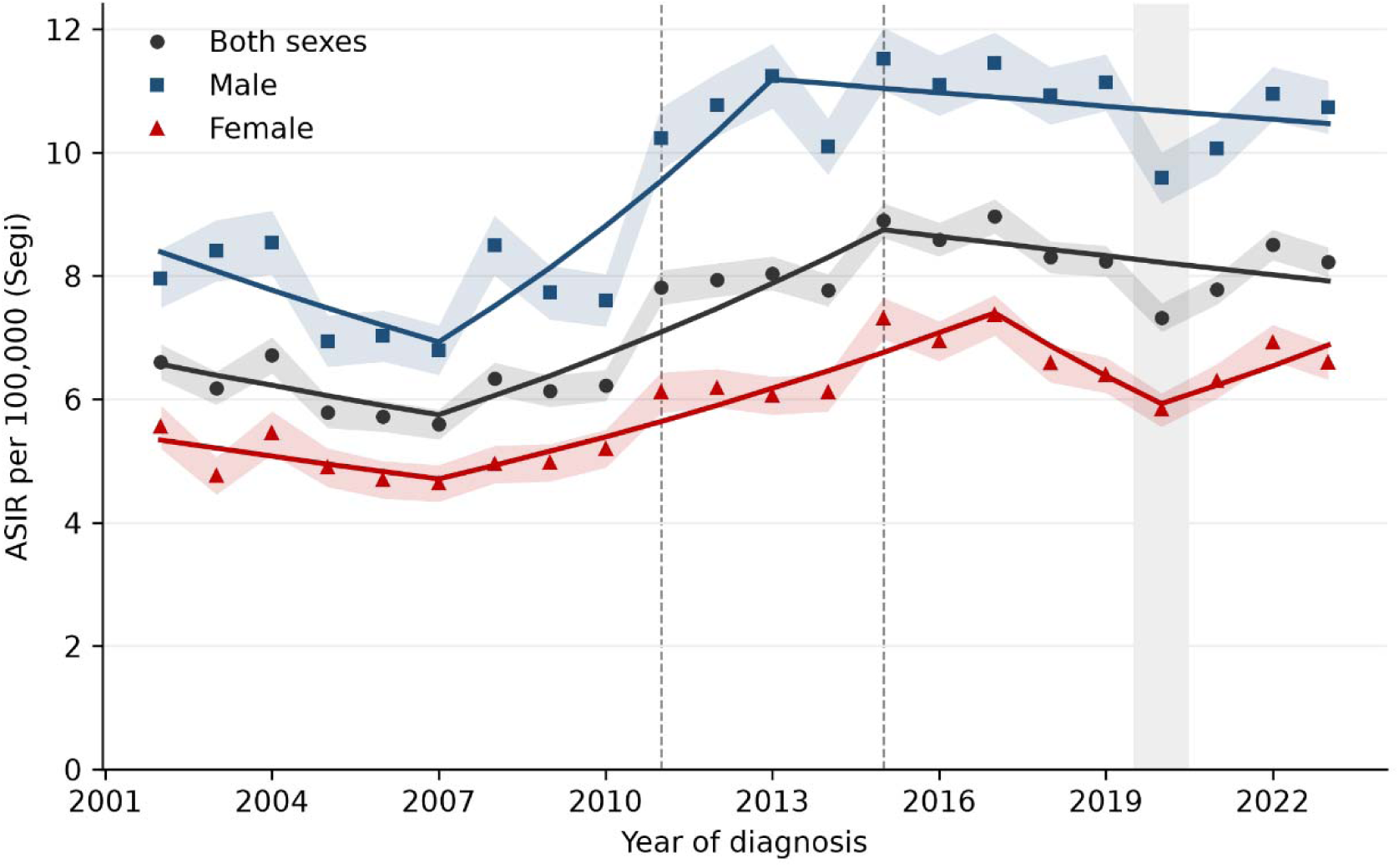
Age-standardised incidence rate (ASIR) of colorectal cancer per 100,000, both sexes and by sex, South Africa, 2002–2023. Rates are standardised to the Segi world population. Shaded bands: 95% confidence intervals. Dashed lines: 2011 (mandatory private-laboratory reporting) and 2015 (change in SANCR case-finding). Grey band: 2020 (COVID-19). Points: observed rates; lines: Joinpoint fits (Table 1).

**Table 1.**
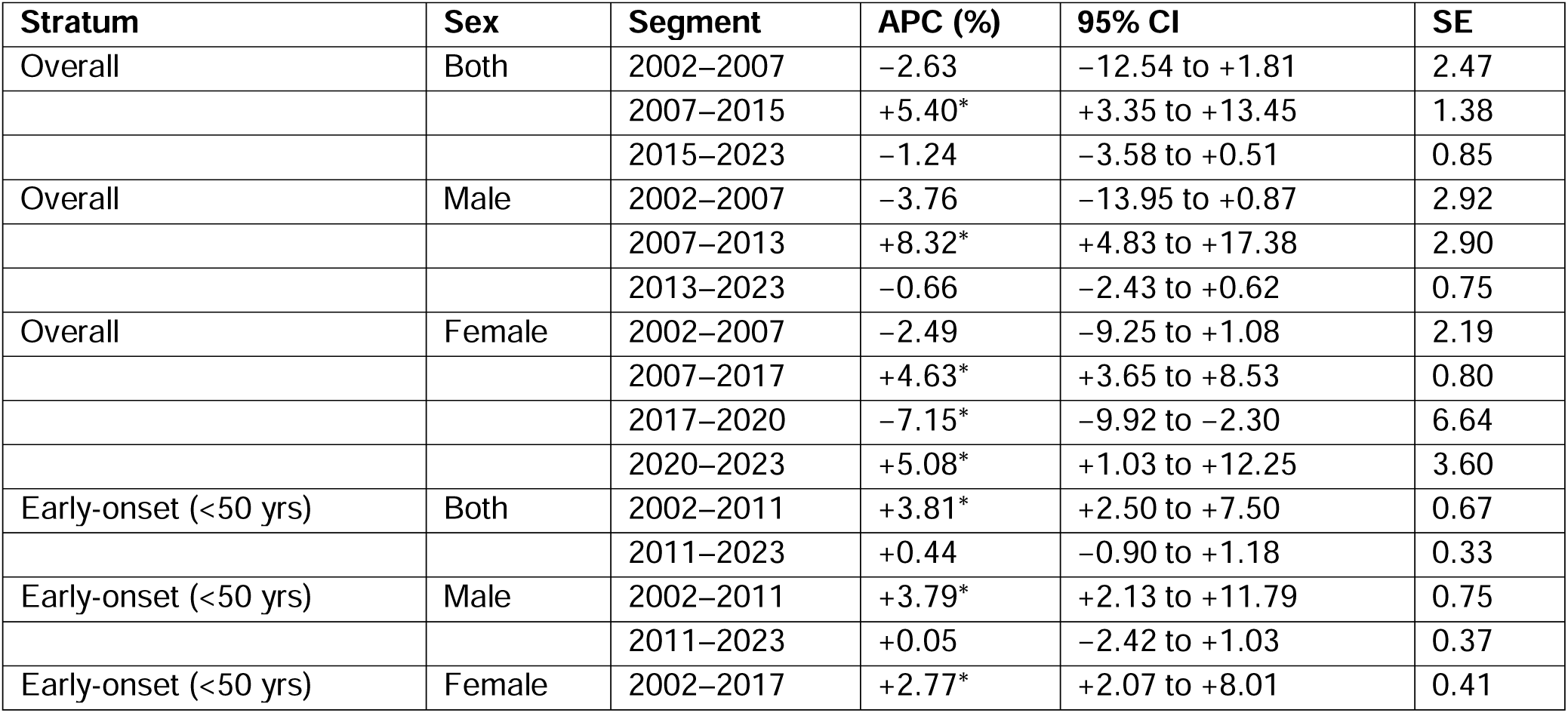
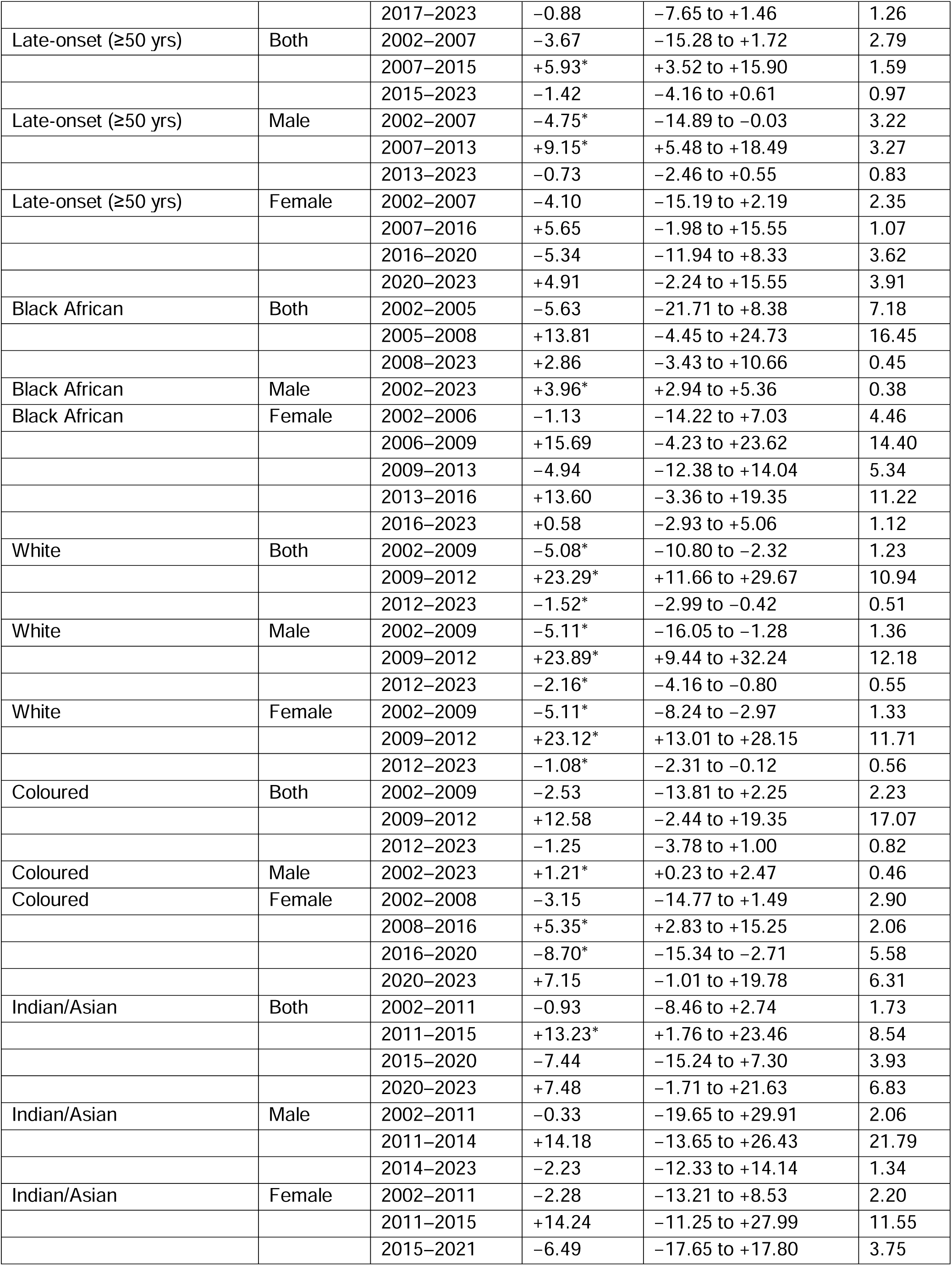
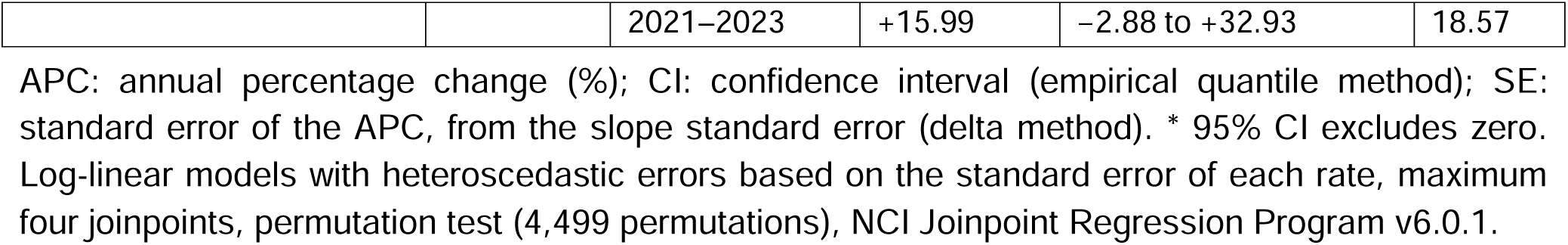
Joinpoint regression APC summary by stratum and sex, SANCR 2002–2023.

### 3.2 Sex-Stratified Incidence

Male ASIR was consistently higher than female ASIR throughout the study period (Figure 1), with a male-to-female ratio between 1.4 and 1.9. Male ASIR increased from 7.96 per 100,000 in 2002 (95% CI 7.49–8.43) to 10.74 in 2023 (10.31–11.17), with a peak of 11.52 in 2015 (11.01–12.03). Female ASIR rose from 5.55 (5.20–5.90) to 6.60 (6.32–6.88), with a peak of 7.36 in 2017 (7.03–7.69).

In males, Joinpoint regression identified two change points, in 2007 and 2013: incidence did not change significantly from 2002 to 2007 (APC −3.76%, 95% CI −13.95 to 0.87), rose from 2007 to 2013 (+8.32%, 4.83 to 17.38) and did not change significantly from 2013 to 2023 (−0.66%, −2.43 to 0.62). The female trend had three change points, in 2007, 2017 and 2020: no significant change from 2002 to 2007 (−2.49%, −9.25 to 1.08), a rise from 2007 to 2017 (+4.63%, 3.65 to 8.53), a decline from 2017 to 2020 (−7.15%, −9.92 to −2.30), which began before the COVID-19 pandemic and ended with the 2020 drop in registrations, and a rise from 2020 to 2023 (+5.08%, 1.03 to 12.25). Annual case counts by sex are shown in Supplementary Figure S1.

### 3.3 Early-Onset versus Late-Onset Colorectal Cancer

Of the 70,467 total cases, 13,195 (18.7%) were diagnosed before age 50 years (early-onset colorectal cancer) and 57,272 (81.3%) at age 50 years or older (late-onset colorectal cancer) (Supplementary Table S1). Truncated ASIR trends by sex are presented in Figure 2.

**Figure 2.**
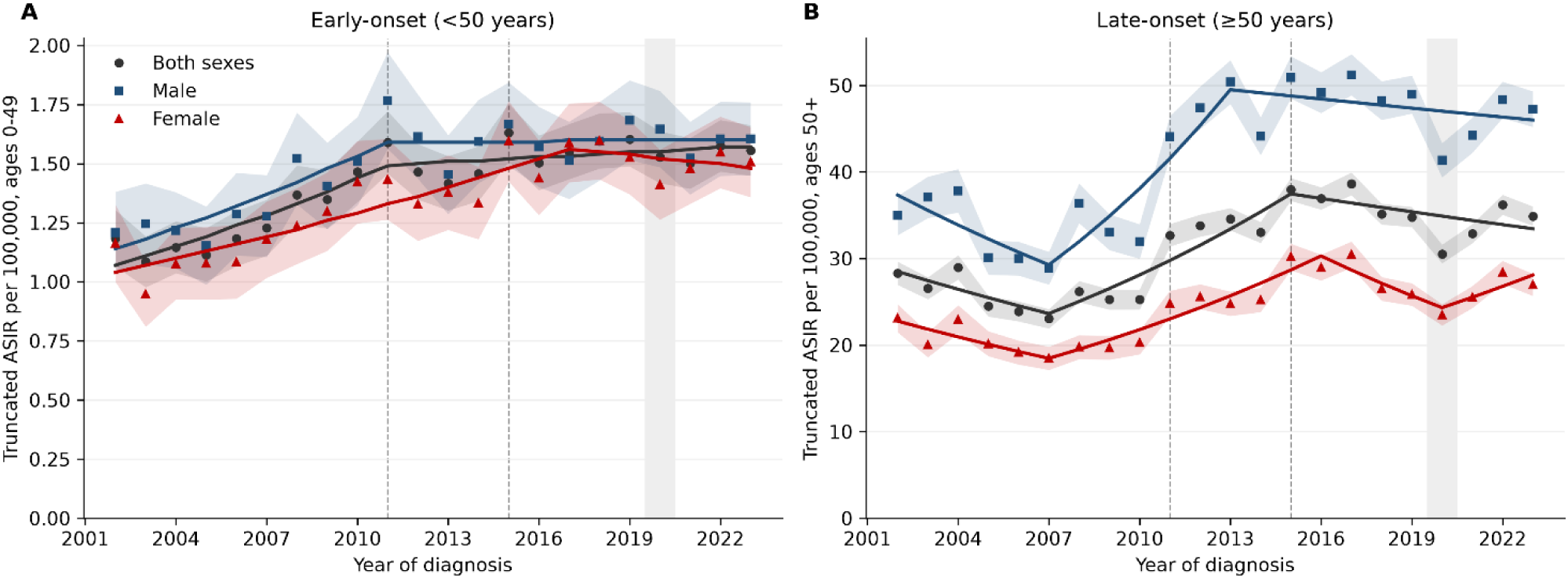
Truncated age-standardised incidence rate (ASIR) per 100,000 of (A) early-onset (<50 years) and (B) late-onset (≥50 years) colorectal cancer, both sexes and by sex, South Africa, 2002–2023. Segi weights for ages 0–49 (A) and 50 and older (B) were rescaled to sum to 100,000. Shaded bands: 95% confidence intervals. Dashed lines: 2011 and 2015; grey band: 2020. The y-axis scales of panels A and B differ. Points: observed rates; lines: Joinpoint fits (Table 1).

The early onset colorectal cancer truncated ASIR rose from 1.18 per 100,000 in 2002 (95% CI 1.07–1.30) to a peak of 1.63 in 2015, and then fell to 1.56 (1.45–1.66) in 2023. Joinpoint regression (combined sex) identified a single change point in 2011 (95% CI 2007–2016): early-onset incidence rose from 2002 to 2011 (APC +3.81%, 95% CI 2.50 to 7.50) and did not change significantly from

2011 to 2023 (+0.44%, −0.90 to 1.18) (Table 1). In males, the change point was also in 2011 (2002–2011: +3.79%, 2.13 to 11.79; 2011–2023: +0.05%, −2.42 to 1.03). In females, incidence rose from 2002 to 2017 (+2.77%, 2.07 to 8.01) and did not change significantly from 2017 to 2023 (−0.88%, −7.65 to 1.46). Over 2011–2023, the early-onset APC was +0.3% (−0.3 to 0.9). No age band under 50 showed a significant change after 2011 (20–29 years +1.0%, −0.8 to 2.8; 30–39 years −0.1%, −0.7 to 0.6; 40–49 years +0.4%, −0.5 to 1.2).

The late onset colorectal cancer truncated ASIR increased from 28.30 per 100,000 in 2002 to a peak of 38.64 in 2017, then fell to 34.91 in 2023. The combined-sex late-onset series had the same structure as the overall series, with change points in 2007 and 2015: no significant change from 2002 to 2007 (APC −3.67%, 95% CI −15.28 to 1.72), a rise from 2007 to 2015 (+5.93%, 3.52 to 15.90) and no significant change from 2015 to 2023 (−1.42%, −4.16 to 0.61). In males, late-onset incidence fell from 2002 to 2007 (−4.75%, −14.89 to −0.03), rose from 2007 to 2013 (+9.15%, 5.48 to 18.49) and did not change significantly from 2013 to 2023 (−0.73%, −2.46 to 0.55). The female late-onset series had change points in 2007, 2016 and 2020, and none of its four segments changed significantly (Table 1). Over 2011–2023, the late-onset APC was 0.0% (95% CI −1.0 to 1.0) (Table 2).

**Table 2.**
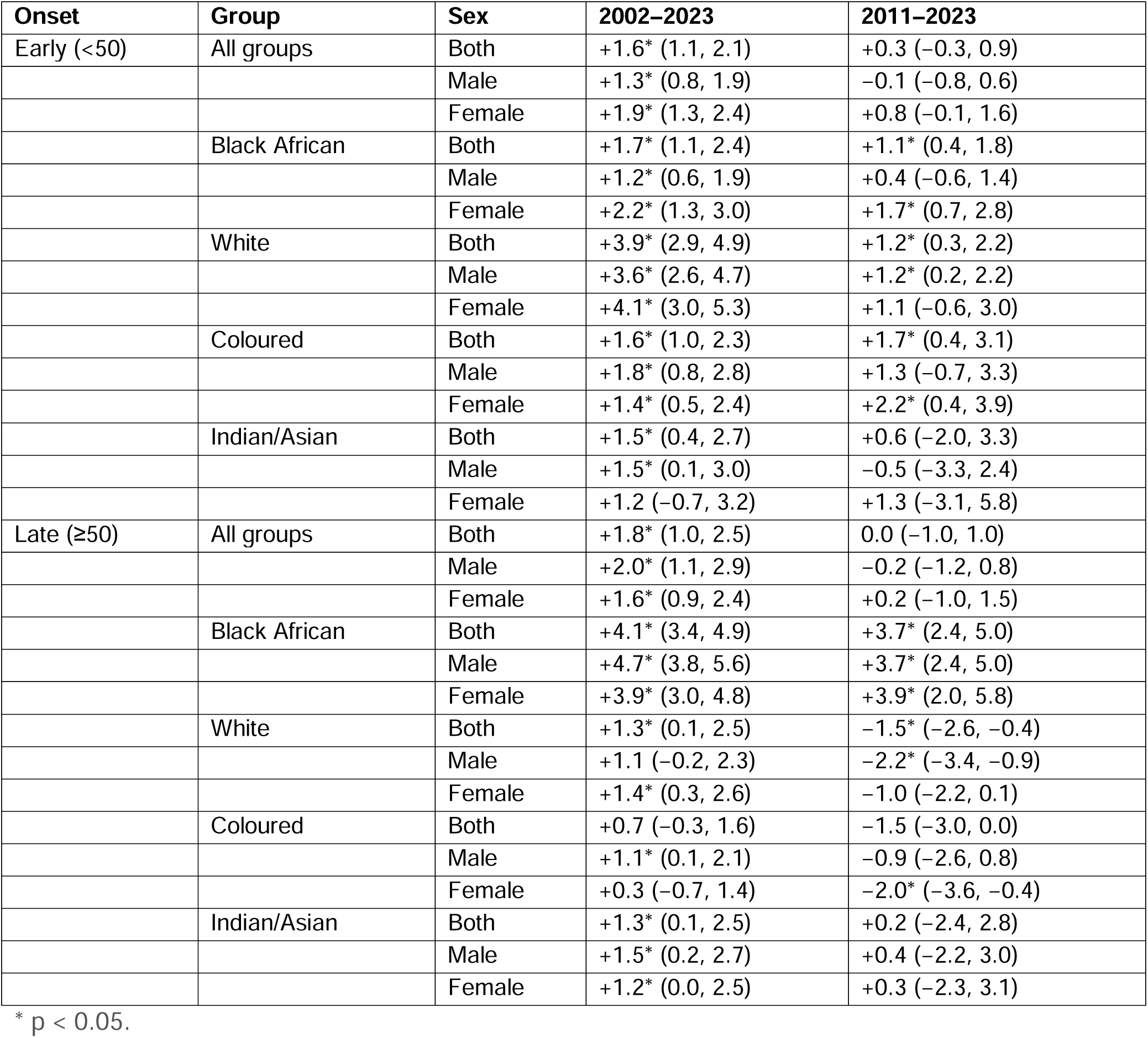
Annual percentage change in colorectal cancer incidence by population group and sex, SANCR 2002–2023.

| **Onset** | **Group** | **Sex** | **2002–2023** | **2011–2023** |
|---|---|---|---|---|
| Early (<50) | All groups | Both | +1.6* (1.1, 2.1) | +0.3 (−0.3, 0.9) |
|  |  | Male | +1.3* (0.8, 1.9) | −0.1 (−0.8, 0.6) |
|  |  | Female | +1.9* (1.3, 2.4) | +0.8 (−0.1, 1.6) |
|  | Black African | Both | +1.7* (1.1, 2.4) | +1.1* (0.4, 1.8) |
|  |  | Male | +1.2* (0.6, 1.9) | +0.4 (−0.6, 1.4) |
|  |  | Female | +2.2* (1.3, 3.0) | +1.7* (0.7, 2.8) |
|  | White | Both | +3.9* (2.9, 4.9) | +1.2* (0.3, 2.2) |
|  |  | Male | +3.6* (2.6, 4.7) | +1.2* (0.2, 2.2) |
|  |  | Female | +4.1* (3.0, 5.3) | +1.1 (−0.6, 3.0) |
|  | Coloured | Both | +1.6* (1.0, 2.3) | +1.7* (0.4, 3.1) |
|  |  | Male | +1.8* (0.8, 2.8) | +1.3 (−0.7, 3.3) |
|  |  | Female | +1.4* (0.5, 2.4) | +2.2* (0.4, 3.9) |
|  | Indian/Asian | Both | +1.5* (0.4, 2.7) | +0.6 (−2.0, 3.3) |
|  |  | Male | +1.5* (0.1, 3.0) | −0.5 (−3.3, 2.4) |
|  |  | Female | +1.2 (−0.7, 3.2) | +1.3 (−3.1, 5.8) |
| Late (≥50) | All groups | Both | +1.8* (1.0, 2.5) | 0.0 (−1.0, 1.0) |
|  |  | Male | +2.0* (1.1, 2.9) | −0.2 (−1.2, 0.8) |
|  |  | Female | +1.6* (0.9, 2.4) | +0.2 (−1.0, 1.5) |
|  | Black African | Both | +4.1* (3.4, 4.9) | +3.7* (2.4, 5.0) |
|  |  | Male | +4.7* (3.8, 5.6) | +3.7* (2.4, 5.0) |
|  |  | Female | +3.9* (3.0, 4.8) | +3.9* (2.0, 5.8) |
|  | White | Both | +1.3* (0.1, 2.5) | −1.5* (−2.6, −0.4) |
|  |  | Male | +1.1 (−0.2, 2.3) | −2.2* (−3.4, −0.9) |
|  |  | Female | +1.4* (0.3, 2.6) | −1.0 (−2.2, 0.1) |
|  | Coloured | Both | +0.7 (−0.3, 1.6) | −1.5 (−3.0, 0.0) |
|  |  | Male | +1.1* (0.1, 2.1) | −0.9 (−2.6, 0.8) |
|  |  | Female | +0.3 (−0.7, 1.4) | −2.0* (−3.6, −0.4) |
|  | Indian/Asian | Both | +1.3* (0.1, 2.5) | +0.2 (−2.4, 2.8) |
|  |  | Male | +1.5* (0.2, 2.7) | +0.4 (−2.2, 3.0) |
|  |  | Female | +1.2* (0.0, 2.5) | +0.3 (−2.3, 3.1) |

\* p < 0.05.

### 3.4 Population Group–Specific Incidence Trends

Substantial differences in colorectal cancer ASIR were observed across South Africa’s four population groups throughout the study period (Figure 3). Averaged over 2002–2023, White South Africans had the highest ASIR (18.4 per 100,000), followed by Indian/Asian (12.4), Coloured (10 9) and Black African (3.0) South Africans; Coloured rates were higher than Indian/Asian rates in 2002, 2004, 2011 and 2012. Despite the lowest per capita rates, Black Africans contributed the larg st absolute case volume owing to their larger population. Early-onset incidence rose in all four population groups over 2002–2023, most steeply in White South Africans (Figure 4A; Table 2).

**Figure 3.**
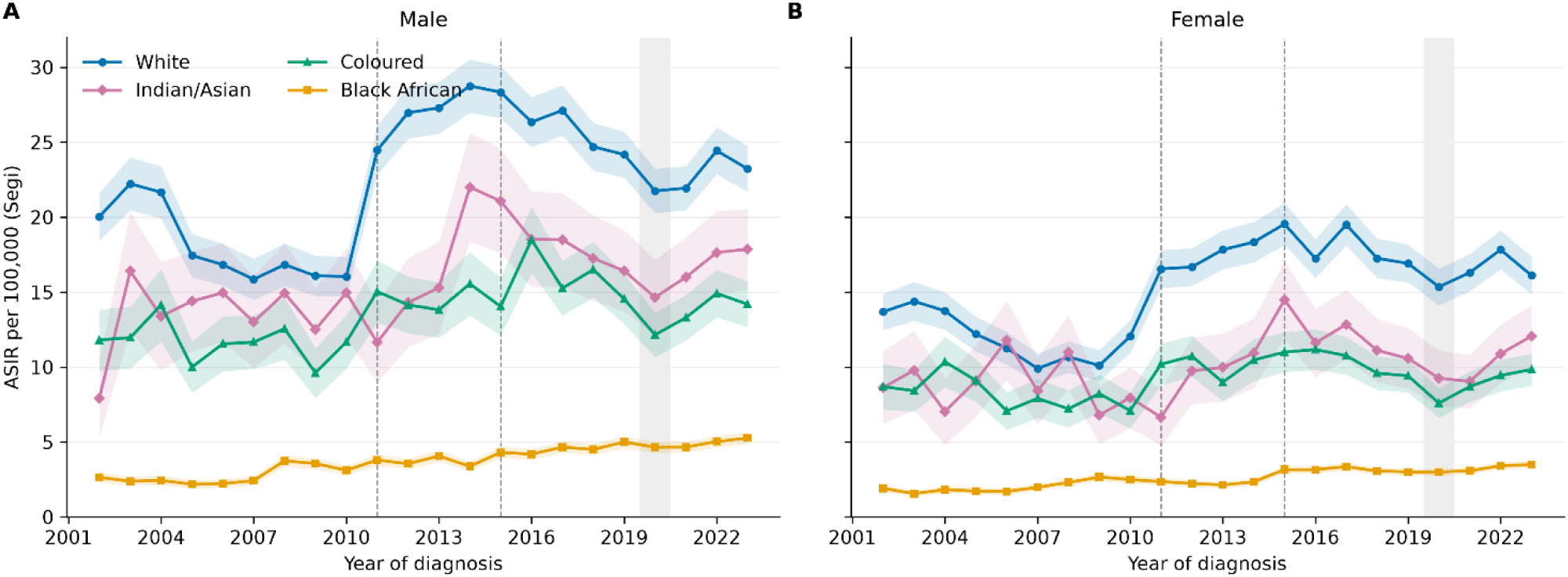
Age-standardised incidence rate (ASIR) of colorectal cancer per 100,000 by population group, (A) males and (B) females, South Africa, 2002–2023. Both panels use the same y-axis scale. Cases with unrecorded population group are excluded from group-specific rates. Shaded bands: 95% confidence intervals. Dashed lines: 2011 and 2015; grey band: 2020.

**Figure 4.**
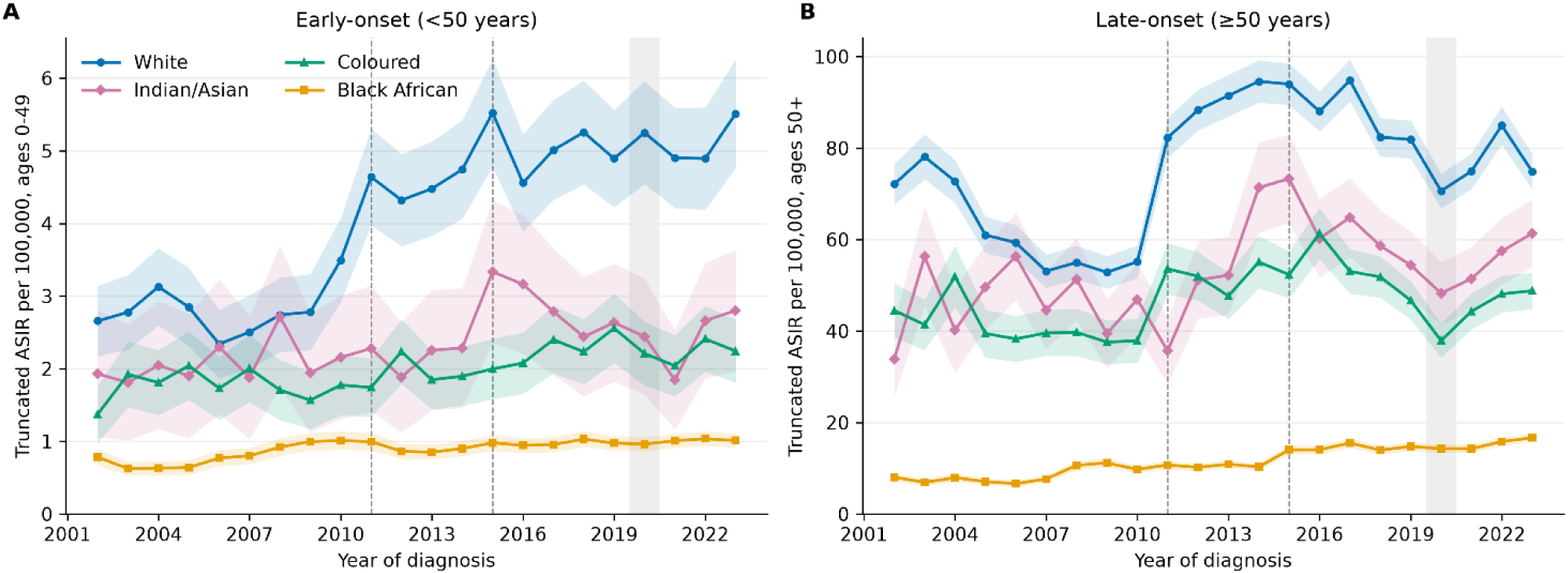
Truncated age-standardised incidence rate per 100,000 of (A) early-onset (<50 years) and (B) late-onset (≥50 years) colorectal cancer by population group, both sexes, South Africa, 2002–2023. Cases with unrecorded population group are excluded. Shaded bands: 95% confidence intervals. Dashed lines: 2011 and 2015; grey band: 2020. The y-axis scales of panels A and B differ.

#### White population

ASIR ranged from 12.63 per 100,000 (2007) to 23.22 per 100,000 (2015). Joinpoint regression identified a significant decline (2002–2009; APC −5.08; p < 0.05), a very sharp and significant rise (2009–2012; APC +23.29; p < 0.05), and a subsequent significant decline (2012–2023; APC −1.52; p < 0.05). The abrupt 2009–2012 increase in all White sub-series (male APC +23.89, female +23.12) is consistent with a registration artefact rather than a true incidence surge, reflecting improved capture of private-laboratory diagnoses. The early-onset APC in White South Africans was +3.9% (95% CI 2.9 to 4.9) during the 2002–2023 period (Figure 4A). The late-onset APC was +1.3% (0.1 to 2.5) over 2002–2023. After 2011, early-onset incidence rose by 1.2% per year (0.3 to 2.2) and late-onset incidence fell by 1.5% per year (−2.6 to −0.4). Early-onset ASIR in White South Africans rose from 2.74 per 100,000 in 2008 to 4.64 in 2011 and was 5.51 in 2023, so much of the 2002–2023 early-onset APC of +3.9% falls in the years of the reporting change.

#### Indian/Asian population

ASIR was relatively stable from 2002 to 2011 (APC −0.93), rose sharply from 2011 to 2015 (APC +13.23; p < 0.05), then declined non-significantly. The ASIR peaked at 17.33 per 100,000 in 2015 and was 14.51 in 2023. The early-onset APC was +1.5% (95% CI 0.4 to 2.7) over 2002–2023 (Figure 4A). The late-onset APC was +1.3% (0.1 to 2.5) over 2002–2023. After 2011, neither early-onset (+0.6%, −2.0 to 3.3) nor late-onset (+0.2%, −2.4 to 2.8) incidence changed significantly.

#### Coloured population

Combined-sex ASIR trends did not reach statistical significance in individ al segments. Male ASIR showed a monotone significant increase over the full period (APC +1.21; p < 0.05). Females showed a significant rise from 2008 to 2016 (APC +5.35; p < 0.05) and a significant decline from 2016 to 2020 (APC −8.70; p < 0.05). The early-onset APC was +1.6% (95% CI 1.0 to 2.3) over 2002–2023 (Figure 4A). The late-onset APC was +0.7% (−0.3 to 1.6) over 2002–2023. After 2011, early-onset incidence rose by 1.7% per year (0.4 to 3.1) and late-onset incidence did not change significantly (−1.5%, −3.0 to 0.0).

#### Black African population

ASIR increased from 2.23 per 100,000 in 2002 to 4.15 in 2023. The combined-sex Joinpoint analysis showed two early transient segments before a rise from 2008 to 2023 that was not statistically significant (APC +2.86). Male Black African ASIR showed a monotone significant increase over the full period (APC +3.96; p < 0.05), consistent with genuine long-term incidence growth in this group. The early-onset APC was +1.7% (95% CI 1.1 to 2.4) over 2002–2023 (Figure 4A). The late-onset APC was +4.1% (3.4 to 4.9) over 2002–2023 and +3.7% (2.4 to 5.0) over 2011–2023, against +1.7% (1.1 to 2.4) and +1.1% (0.4 to 1.8) for early-onset. Incidence rose in Black African women as well as men (all ages: women +3.4%, 2.7 to 4.2; men +4.0%, 3.2 to 4.7).

### 3.5 Joinpoint Regression Summary

Table 1 summarises the Joinpoint results for all 21 series; * marks APCs whose 95% CI excludes zero. Overall, male, female, late-onset and late-onset male series rose from 2007 to 2013–2017, the period of expanded private-sector reporting, and did not change significantly afterwards, except for females, whose incidence fell from 2017 to 2020 and rose from 2020 to 2023. Early-onset incidence (both sexes and males) rose until 2011 and did not change significantly afterwards. Black African males (APC +3.96%, 95% CI 2.94 to 5.36) and Coloured males (+1.21%, 0.23 to 2.47) had the only increases over the whole period with no change point.

### 3.6 Sensitivity Analyses

**SA1, restricted to 2011–2023:** for all population groups combined, neither early-onset (+0.3% per year, 95% CI −0.3 to 0.9) nor late-onset (0.0%, −1.0 to 1.0) incidence changed significantly (Table 2; Table 3). Within population groups, early-onset incidence rose in White (+1.2%, 0.3 to 2.2), Coloured (+1.7%, 0.4 to 3.1) and Black African (+1.1%, 0.4 to 1.8) South Africans, at a slower rate than over 2002–2023 in White South Africans (+3.9%). Late-onset incidence fell in White South Africans (−1.5%) and rose in Black African South Africans (+3.7%). The rise in early-onset incidence over 2002–2023 therefore owes much to the period of reporting change, and a smaller rise persists within population groups after 2011.

**Table 3.** Poisson models. APC % per year, early-onset / late-onset.

| Group | Primary 2002–2023 | p (early vs late) | SA1 2011–2023 | SA2 excl. 2020 | Level shift at 2011, IRR (95% CI) |
|---|---|---|---|---|---|
| All | +1.5* / +1.8* | 0.267 | +0.3 / 0.0 | +1.5* / +2.0* | 1.35 (1.27–1.42) |
| Black African | +1.6* / +4.3* | <0.001 | +1.0 / +3.7* | +1.6* / +4.3* | 1.03 (0.95–1.12) |
| Coloured | +1.6* / +0.7* | 0.095 | +1.6* / −1.3* | +1.6* / +1.0* | 1.36 (1.25–1.49) |
| Indian/Asian | +1.2 / +1.4* | 0.776 | +0.3 / +0.5 | +1.3 / +1.6* | 1.14 (0.99–1.33) |
| White | +4.0* / +0.9* | <0.001 | +1.1 / −2.2* | +4.0* / +1.0* | 1.77 (1.65–1.91) |

\* p < 0.05.

**SA2, Exclusion of 2020:** Removal of the 2020 year did not materially alter point estimates or the significance of the year coefficient in any stratum. The COVID-19 pandemic is therefore unlikely to have substantially biased the primary regression estimates, though the 2020 ASIR dip remains an acknowledged artefact of the annual rate series. Excluding 2020, the Poisson year slopes were +1.5% (early-onset) and +2.0% (late-onset) per year, against +1.5% and +1.8% in the primary model (Table 3).

**SA3, testing the 2011 mandate:** A level shift at 2011 added to the primary model gave a rate ratio of 1.35 (95% CI 1.27–1.42) for all groups (Table 3). The shift was largest in White (1.77, 1.65–1.91) and Coloured (1.36, 1.25–1.49) South Africans and absent in Black African South Africans (1.03, 0.95–1.12). With the step included, the early-onset year slope was −0.3% per year and the late-onset slope 0.0%.

### 3.7 Early- and late-onset mortality

Of 40,036 colorectal cancer deaths in 2002–2019, 5,988 (15.0%) were under age 50. Early-onset ASMR was 0.74 per 100,000 in 2002 and 0.81 in 2019, and late-onset ASMR 21.50 and 24.89 (Figure 5). Over 2002–2019, early-onset mortality rose by 1.2% per year (95% CI 0.5 to 2.0) and late-onset mortality by 0.9% (0.3 to 1.4). After 2011, neither changed significantly (early-onset +0.6%, −1.3 to 2.6; late-onset +1.1%, −0.2 to 2.5). Deaths fell in 2020 in both onset groups and had not returned to the 2019 level by 2022. By population group, late-onset mortality rose most in Black African South Africans (+5.1% per year over 2002–2019, +5.0% over 2011–2019) (Supplementary Table S3; Supplementary Figure S4). Early-onset mortality rose in White (+2.9%, 1.6 to 4.2) and Coloured (+3.8%, 2.4 to 5.2) South Africans, while late-onset mortality did not change in White South Africans (+0.1%, −0.5 to 0.7). Group-specific mortality trends over 2002–2019 are affected by the fall in deaths with unrecorded population group, from 20–24% in 2002–2010 to 8–14% thereafter (Supplementary Figure S2). Excluding these deaths, as in earlier analyses, gives an all-ages APC of +2.2% instead of +0.9%.

**Figure 5.**
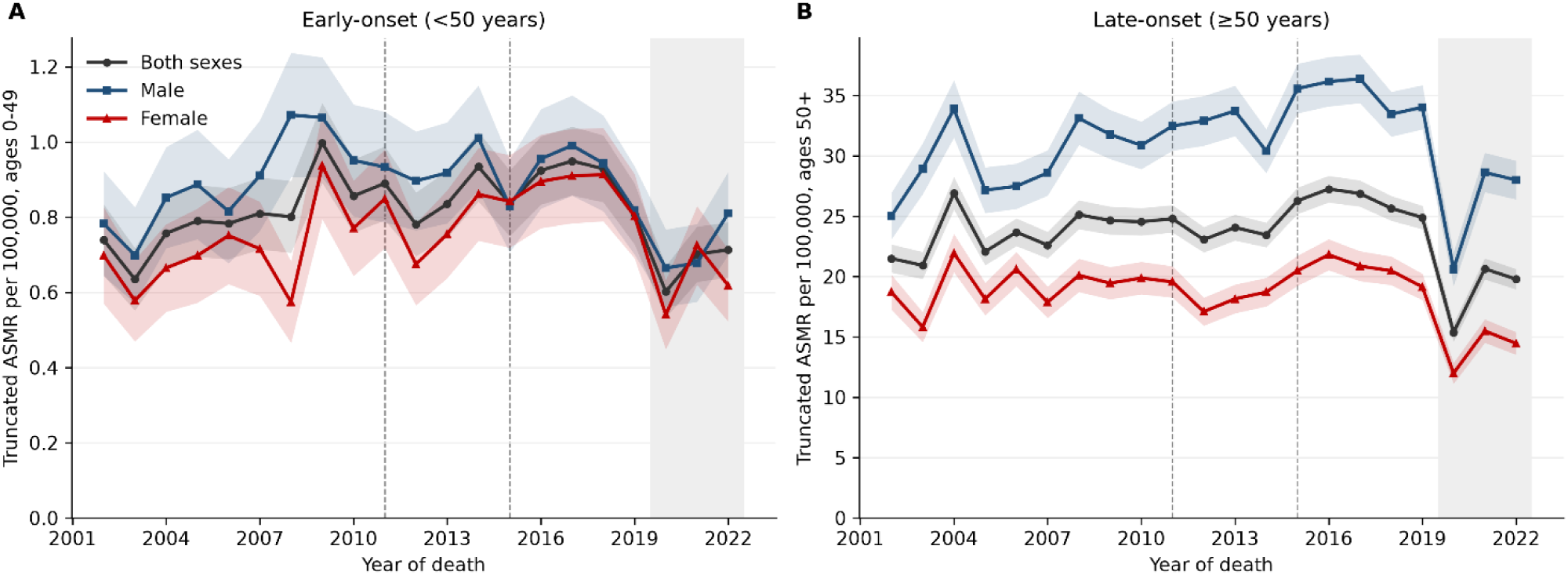
Truncated age-standardised mortality rate (ASMR) per 100,000 of (A) early-onset (<50 years) and (B) late-onset (≥50 years) colorectal cancer, both sexes and by sex, South Africa, 2002–2022. Underlying cause of death C18–C20, Stats SA death records; deaths with unrecorded population group are included. Shaded bands: 95% confidence intervals. Grey band: 2020–2022, outside the primary mortality window (2002–2019).

### 3.8 Mortality-to-incidence ratio

The mean MIR in 2011–2019 was 0.57 for early-onset and 0.71 for late-onset disease (Figure 6). For early-onset disease, it was 0.68 in Black African, 0.56 in Coloured, 0.44 in Indian/Asian and 0.29 in White South Africans; for late-onset disease 0.92, 0.62, 0.62 and 0.53.

**Figure 6.**
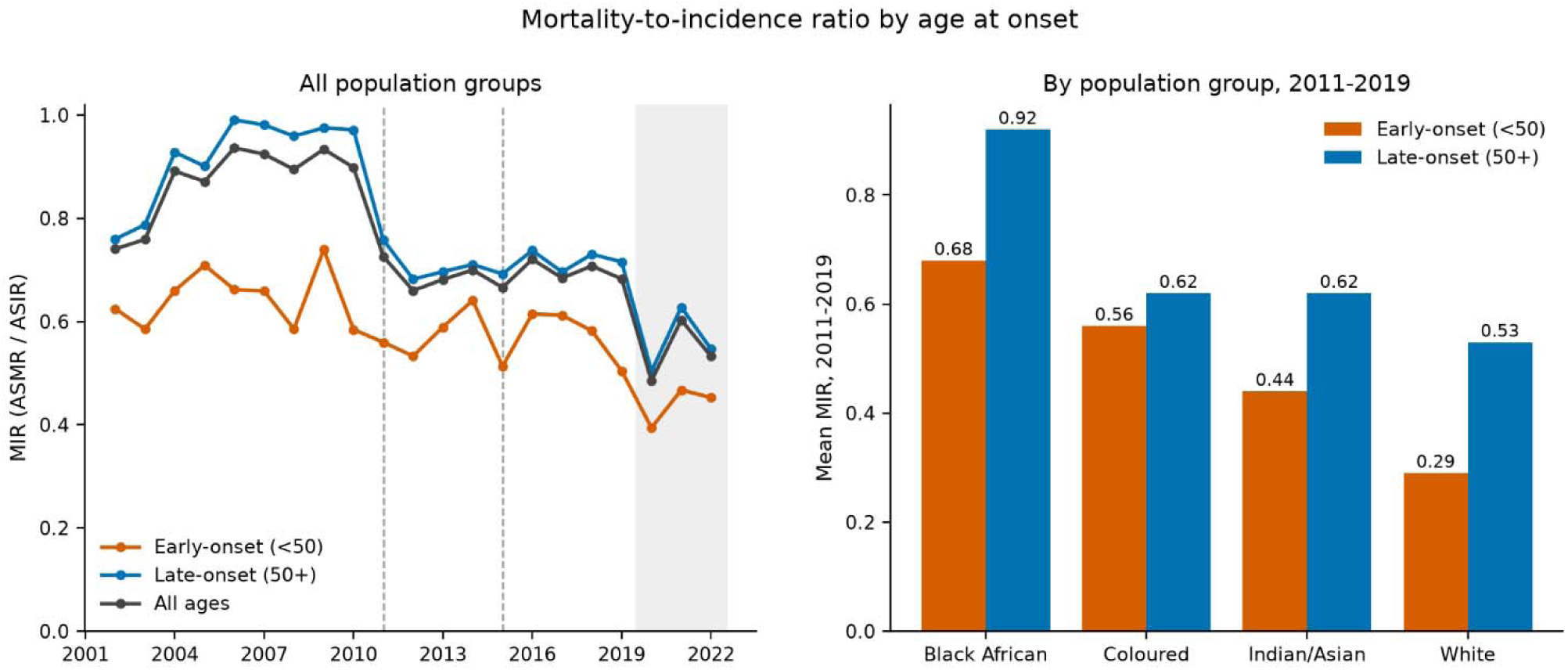
Mortality-to-incidence ratio (MIR = ASMR / ASIR) of colorectal cancer by age at onset. (A) Annual MIR, all population groups, 2002–2022. (B) Mean MIR in 2011–2019 by population group and age at onset. The MIR is an ecological ratio and does not measure survival.

### 3.9 Provincial mortality

Of 33,319 deaths in 2006–2019, 33,064 were assigned to a province (Figure 7; Supplementary Table S2). Early-onset ASMR ranged from 0.57 per 100,000 in Mpumalanga to 1.39 in the Northern Cape (2.4-fold), and late-onset ASMR ranged from 11.6 in Limpopo to 45.5 in the Western Cape (3.9-fold). The Western Cape and Northern Cape were among the three highest provinces in both onset groups. Gauteng ranked second for late-onset mortality (31.6) but was below the national rate for early-onset mortality (0.84). The gradient was steeper for late-onset disease. Provincial rates are not adjusted for population group.

**Figure 7.**
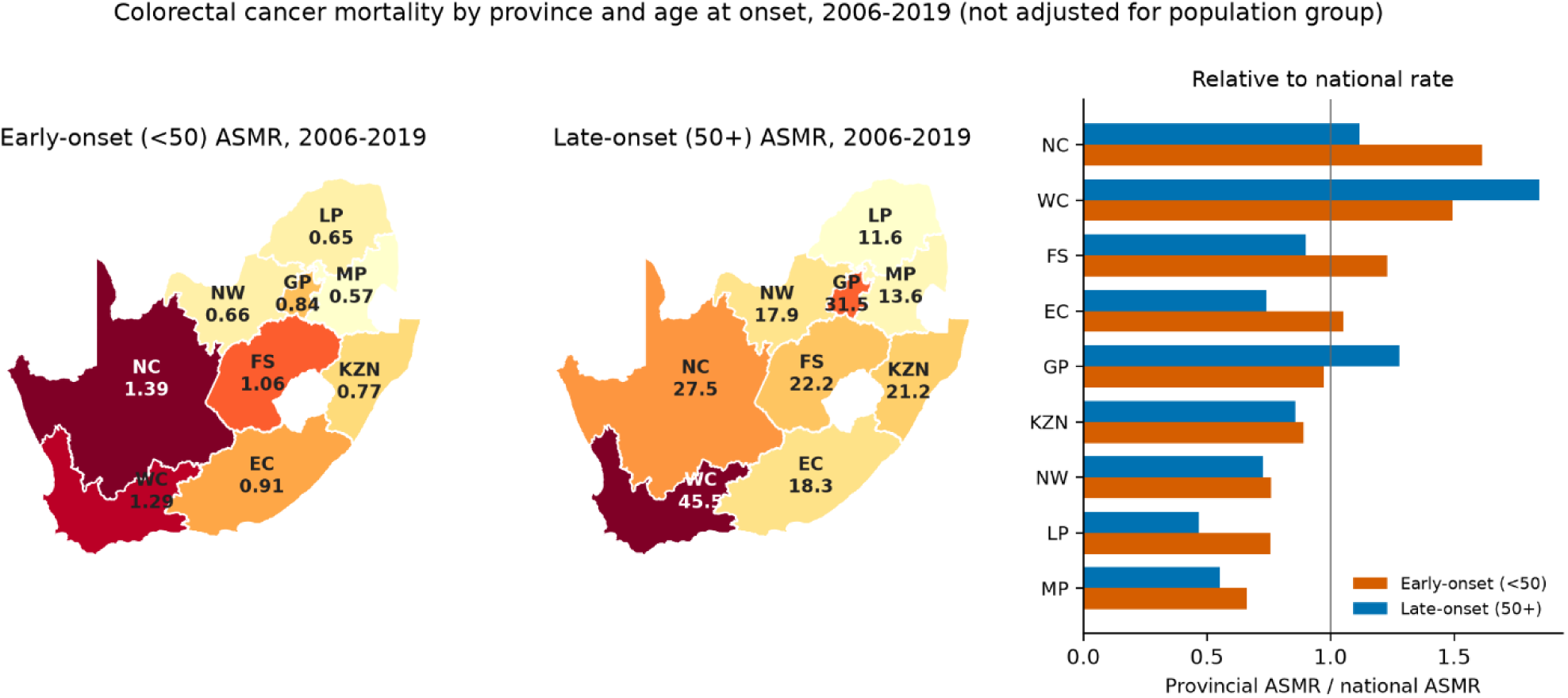
Truncated age-standardised colorectal cancer mortality rates per 100,000 by province and age at onset, South Africa, 2006–2019: early-onset (<50 years) and late-onset (≥50 years) maps, and each province’s rate relative to the national rate. Province of residence, or province of death where residence was missing. Rates are not adjusted for population group. Provinces: EC Eastern Cape, FS Free State, GP Gauteng, KZN KwaZulu-Natal, LP Limpopo, MP Mpumalanga, NC Northern Cape, NW North West, WC Western Cape.

### 3.10 Reporting sector

Early-onset cases were less often diagnosed in private laboratories than late-onset cases in every population group except Indian/Asian: Black African 37% vs 42%, Coloured 39% vs 50%, White 70% vs 79%, Indian/Asian 71% vs 70%.

## 4. Discussion

To our knowledge, this is the first national study in South Africa to assess changes in colorectal cancer incidence and mortality by age at onset. Similar to previous studies using SANCR data^4^, we found that incidence was highest in White South Africans, followed by Indian/Asian, Coloured and Black African South Africans.

Falls in incidence in White South Africans from 2002 to 2009 could reflect drops in case reporting from private-sector laboratories from 2005–2007, as previously reported^4^. Legislative changes in 2011 led to mandatory reporting by private-sector laboratories^9^, which likely explains the abrupt increase in incidence in the 2009–2012 period. SANCR data on White South Africans are likely more sensitive to changes in private sector reporting, given that 72% have private health care insurance compared to 10% of the Black South African population^12^. Extending previous SANCR studies^4^, we found that in White South Africans, overall and late-onset incidence fell from 2012 to 2023 (APC −1.4% and −2.0%), while early-onset incidence rose (+1.3% per year, 95% CI 0.2 to 2.4). The APC of +3.9% over 2002–2023 includes the increase in registrations that followed mandatory private-laboratory reporting and overstates the recent rise. At the post-2012 rate, early-onset incidence would take more than 50 years to double. This pattern is similar to what has been reported in other countries^3^.

In contrast, although overall colorectal cancer incidence is increasing fastest in Black South Africans, confirming a previous study^4^, most of this is being driven by late-onset colorectal cancer. In Black African males, the all-ages APC was +3.96% (95% CI 2.94 to 5.36) over 2002–2023 (Table 1), compared with 4.3% for 2002–2014 in the previous SANCR report^4^. Incidence also rose in Black African females (+3.4% per year, 95% CI 2.7 to 4.2), similar to the 3.4% reported for 2002–2014^4^. The late-onset APC in Black African males was +4.7% (3.8 to 5.6) over 2002–2023 and +3.7% (2.4 to 5.0) over 2011–2023. If the 2011–2023 rate of increase in Black African South Africans (+3.1% per year, 95% CI 2.0 to 4.2) continued, incidence in this group would double in about 23 years; at the 2002–2023 rate (+3.6%, 2.9 to 4.2) it would double in about 20 years.

We observed a modest increase in incidence of early-onset colorectal cancer in Black Africans, with an APC of 1.7%. Modest increases in early-onset colorectal cancer incidence were also observed in coloureds (APC 1.6%) and Indian/Asians (APC 1.5%).

Mortality data give a check on registration. Early-onset mortality rose by 1.2% per year over 2002– 2019, close to the early-onset incidence trend, and did not change significantly after 2011. In Black African South Africans late-onset mortality rose faster than late-onset incidence (5.1% vs 4.1% per year), and the MIR was highest (0.92 for late-onset disease in 2011–2019), consistent with later diagnosis or poorer access to treatment. Early-onset MIR was lowest in White South Africans (0.29). The Western Cape and Northern Cape were among the three provinces with the highest mortality in both onset groups.

Joinpoint regression placed the only change point in early-onset incidence in 2011 (95% CI 2007– 2016), the year private-laboratory reporting became mandatory. For all population groups combined, early-onset incidence did not change after 2011 (Joinpoint APC +0.44% per year, 95% CI −0.90 to 1.18; period APC +0.3%, −0.3 to 0.9), yet it rose within the White, Coloured and Black African groups (+1.1% to +1.7% per year). The pooled rate is a population-weighted average of the group rates. Between 2011 and 2023, the White share of the population under 50, the group with the highest early-onset rate, fell from 9.0% to 7.5%, and the Black African share, the group with the lowest rate, rose from 79.5% to 81.1%. This change in population composition pulls down the pooled trend, so group-specific APCs better capture change within groups. These post-2011 increases are smaller than the 2002–2023 estimates, which include the 2009–2012 reporting step, and are similar in direction to the increases reported in many high-income countries^3^.

In Coloured South Africans, early-onset incidence rose after 2011 (+1.7% per year, 95% CI 0.4 to 3.1) while late-onset incidence did not change significantly (−1.5%, −3.0 to 0.0). Early-onset mortality in this group rose by 3.8% per year over 2002–2019 (2.4 to 5.2) and by 6.0% per year over 2011–2019 (1.9 to 10.3), the latter period being less affected by the fall in deaths with unrecorded population group. In Indian/Asian South Africans, incidence rose sharply from 2011 to 2015 (APC +13.2%) and then declined, and neither early-nor late-onset incidence changed significantly over 2011–2023. This group makes up 2.6% of the population, and its confidence intervals are wide.

Early-onset cases made up 31.7% of cases in Black African patients and 10.9% in White patients, and the median age at diagnosis was 57 and 67 years (Supplementary Table S1). The Black African population is younger, so a larger share of its cases is expected before age 50 even at similar age-specific risk, and early-onset rates per 100,000 remained lowest in Black African South Africans throughout (Figure 4A).

This study covers all histologically confirmed colorectal cancers registered nationally over 22 years and adds national mortality over 18 years. Records with unrecorded population group were kept in the overall, sex and onset rates, which removed an upward bias in the mortality trends of earlier analyses. The effect of reporting changes was assessed with a post-2011 window and with level-shift models.

This study has limitations. The SANCR is pathology-based: cancers diagnosed on clinical or imaging grounds alone are not registered, and completeness depends on laboratory reporting, which changed in 2005–2007, 2011 and 2015. We assessed these changes with a post-2011 window and level-shift models, but neither can separate a reporting change from a real change in incidence that occurred at the same time. Case counts before 2011 are likely underestimates. The 2020 rates are affected by COVID-19 disruption of health services and registration, and deaths had not returned to the 2019 level by 2022, so mortality trends after 2019 could not be assessed. Population group was unrecorded for 1–5% of cases each year, and for 20–24% of deaths in 2002– 2010 and 8–14% thereafter; the fall in unrecorded deaths biases group-specific mortality trends that start before 2011 upwards. Mortality data rely on the underlying cause on death notifications, whose accuracy may differ between provinces and population groups^15^. The mortality-to-incidence ratio is an ecological ratio and does not measure survival^16^. Provincial mortality rates are not adjusted for population group, and provincial incidence could not be estimated because the SANCR records the reporting laboratory rather than the patient’s residence. Population denominators are Stats SA estimates and are less certain for smaller groups; the Indian/Asian group is small, and its estimates are imprecise.

## 5. Conclusions

Colorectal cancer incidence in South Africa rose over 2002–2023 in both early- and late-onset disease, and much of the rise coincided with the 2011 reporting mandate. After 2011, early-onset incidence rose by about 1–2% per year within the White, Coloured and Black African groups. The largest increases in both incidence and mortality were in late-onset disease in Black African South Africans. Monitoring early-onset trends will depend on stable laboratory reporting and on complete recording of population group on death notifications.

## Abbreviations

AAPC: average annual percentage change;
APC: annual percentage change;
ASIR: age-standardised incidence rate;
ASMR: age-standardised mortality rate;
CI: confidence interval;
IRR: incidence rate ratio;
MIR: mortality-to-incidence ratio;
NCI: National Cancer Institute;
SANCR: South African National Cancer Registry;
Stats SA: Statistics South Africa.

## Consent for publication

Not applicable.

## Data availability

Individual-level SANCR data are available from the National Cancer Registry on reasonable request and with approval from relevant ethics committees. Death records are available from Statistics South Africa.

## Code availability

Analysis code is available upon request.

## Author contributions

WCC, JTB, PCH: conceptualisation; SAM, NA, BN, MIM, TT, JRMMK, CMT, ZM, NM, VN, MN: data curation; WCC: formal analysis; WCC, PCH: methodology; WCC: visualisation; WCC, PCH: writing, original draft; [all authors]: writing, review and editing; WCC, PCH, MM: supervision; MM: funding acquisition.

## Members of the Molecular, Somatic and Infectious Drivers of Colorectal Cancer in Africa (MOSAIC) collaboration

*Authors who are members of the MOSAIC collaboration:*

Wenlong Carl Chen, Jean-Tristan Brandenburg, Philip C Haycock

*Non-author members of the MOSAIC collaboration:*

Lisa Campbell, Dewaldt Engelbrecht, Christopher G. Mathew, Richard M Martin, Brendan Bebington, Vuyo Soldati, Sandra Perdomo, Wendy Stevens, June Fabian, Michele Ramsay, Moses Koobotse, Matthew Suderman, Paul D Yousefi, Thomas Vogl, Mandeep Kaur, Adam Chambers, Helen Winter, Boitumelo Ramasodi, Abel David González-Pérez, Nadine Harran, Jennifer Groverman, Nicole Sheahan, Dr Miguel Vanterpool Héctor, Lisa Ware, Olusegun Isaac Alatise

## Acknowledgements

We gratefully acknowledge the staff of the SANCR, the public and private pathology laboratories that report to the SANCR, Statistics South Africa and the participants and patient representatives at the Wits-Bristol colorectal cancer workshop (30 September to 1 October 2025).

## 7. Supplemental Materials

**Supplementary Figure S1.**
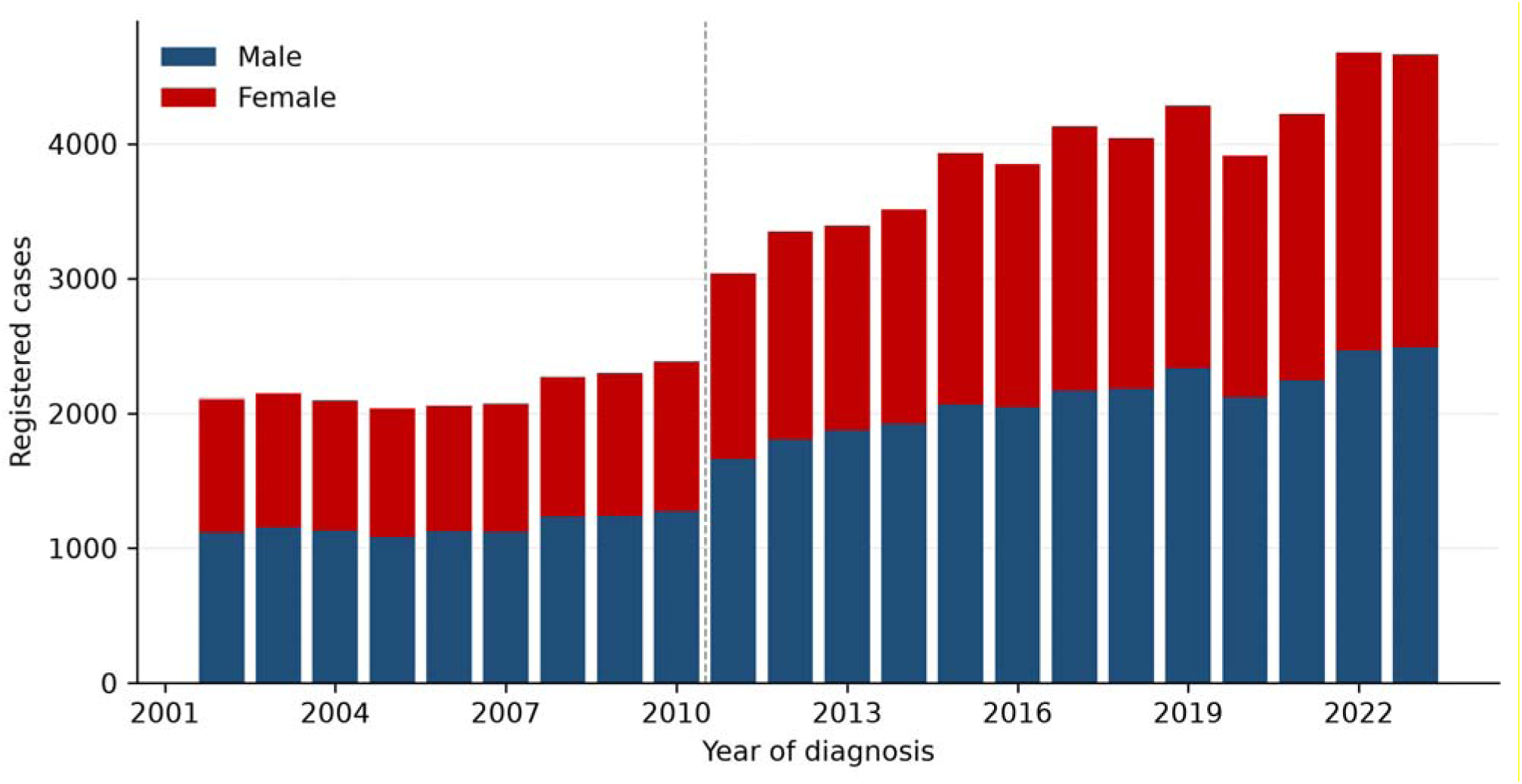
Annual number of registered colorectal cancer cases by sex, South Africa, 2002– 2023. Dashed line: start of mandatory private-laboratory reporting in 2011.

**Supplementary Figure S2.**
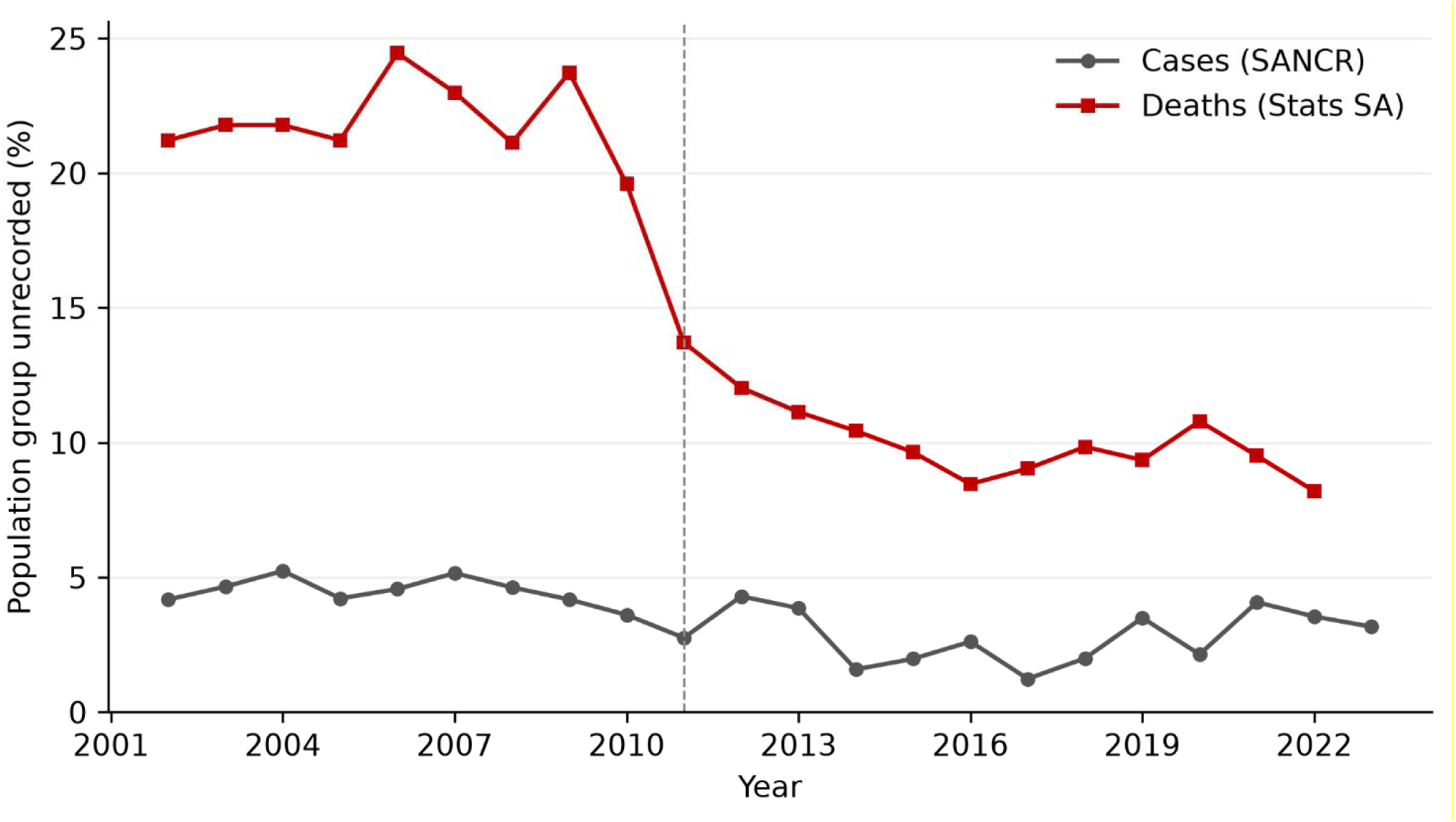
Percentage of colorectal cancer cases (SANCR) and deaths (Stats SA) with unrecorded population group, by year, 2002–2023.

**Supplementary Figure S3.**
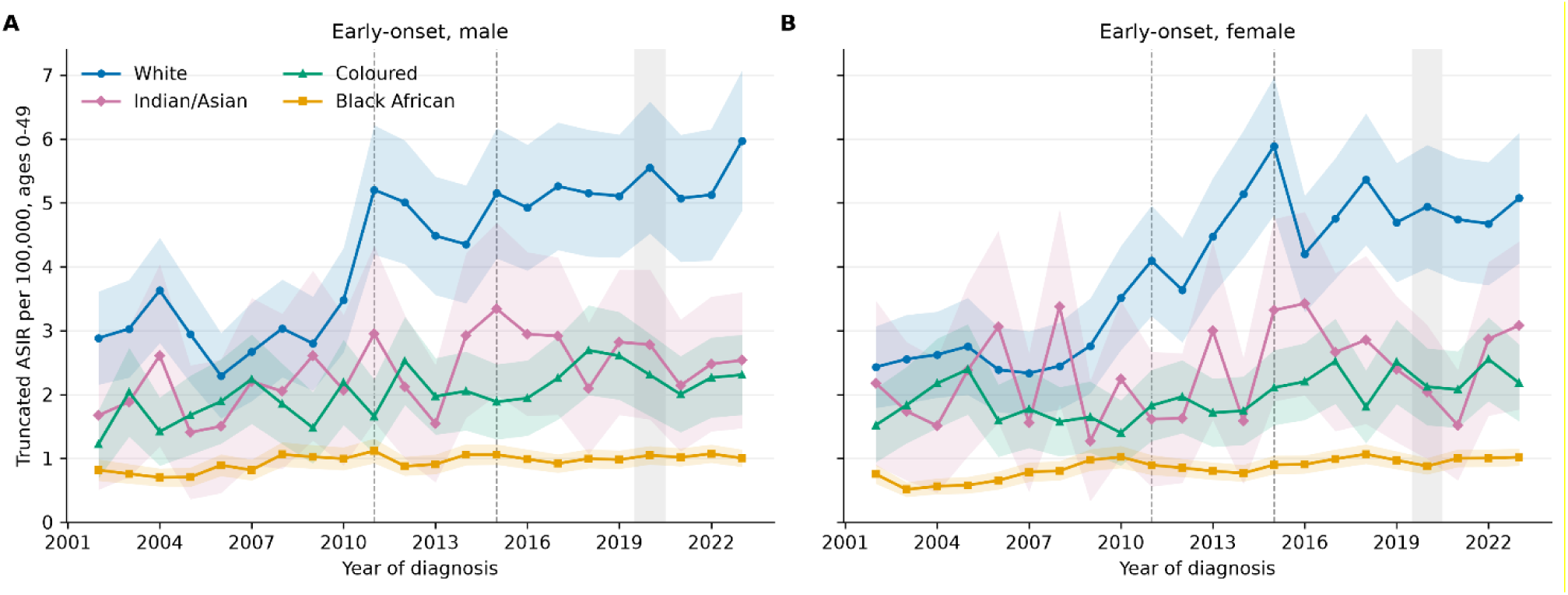
Truncated age-standardised incidence rate of early-onset (<50 years) colorectal cancer per 100,000 by population group, (A) males and (B) females, South Africa, 2002–2023. Shaded bands: 95% confidence intervals. Both panels use the same y-axis scale.

**Supplementary Figure S4.**
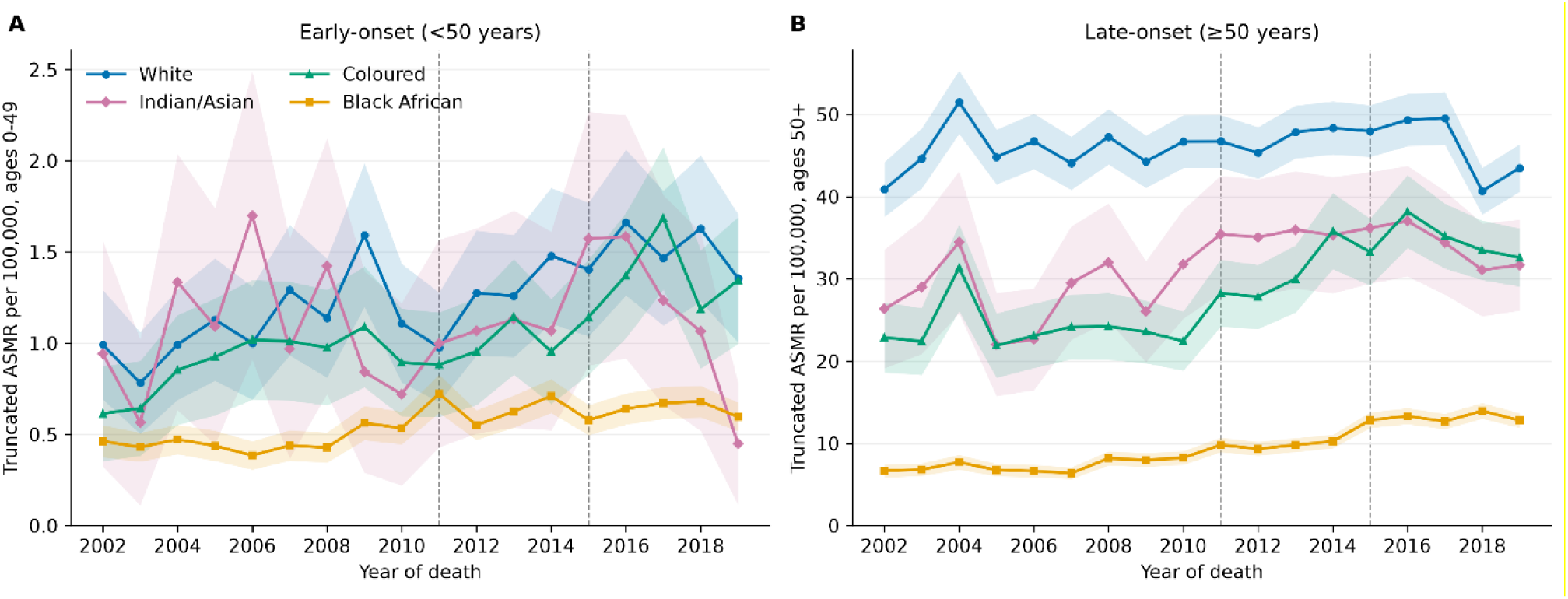
Truncated age-standardised mortality rate per 100,000 of (A) early-onset and (B) late-onset colorectal cancer by population group, South Africa, 2002–2019. Deaths with unrecorded population group are excluded. Shaded bands: 95% confidence intervals.

**Supplementary Table S1.** Number of cases and deaths, percentage early-onset and median age by population group. Incidence 2002–2023, mortality 2002–2019. Population-group rows exclude records with unrecorded group.

| Group | Cases | Early-onset, n (%) | Median age (IQR) | Deaths | Early-onset, n (%) | Median age (IQR) |
|---|---|---|---|---|---|---|
| All | 70,467 | 13,195 (18.7) | 63 (53–72) | 40,036 | 5,988 (15.0) | 66 (56–76) |
| Black African | 20,715 | 6,560 (31.7) | 57 (46–66) | 11,978 | 3,096 (25.8) | 60 (49–70) |
| Coloured | 9,756 | 1,829 (18.7) | 62 (53–71) | 4,383 | 769 (17.5) | 63 (53–72) |
| Indian/Asian | 4,203 | 669 (15.9) | 62 (54–70) | 1,788 | 238 (13.3) | 65 (56–74) |
| White | 33,475 | 3,661 (10.9) | 67 (58–75) | 15,802 | 939 (5.9) | 72 (63–80) |

**Supplementary Table S2.** Truncated age-standardised colorectal cancer mortality rate per 100,000 (95% CI) by province and age at onset, 2006–2019.

| **Province** | **Early-onset deaths** | **Early-onset ASMR** | **Late-onset deaths** | **Late-onset ASMR** |
|---|---|---|---|---|
| Eastern Cape | 548 | 0.91 (0.83–0.99) | 3,156 | 18.31 (17.65–18.96) |
| Free State | 305 | 1.06 (0.94–1.18) | 1,398 | 22.20 (21.04–23.37) |
| Gauteng | 1,298 | 0.84 (0.80–0.89) | 8,121 | 31.55 (30.86–32.24) |
| KwaZulu-Natal | 835 | 0.77 (0.72–0.82) | 4,170 | 21.16 (20.52–21.80) |
| Limpopo | 353 | 0.65 (0.58–0.72) | 1,254 | 11.55 (10.91–12.20) |
| Mpumalanga | 260 | 0.57 (0.50–0.64) | 1,078 | 13.65 (12.83–14.47) |
| North West | 249 | 0.66 (0.57–0.74) | 1,215 | 17.89 (16.88–18.90) |
| Northern Cape | 163 | 1.39 (1.18–1.61) | 738 | 27.51 (25.50–29.51) |
| Western Cape | 923 | 1.29 (1.21–1.37) | 7,000 | 45.53 (44.46–46.60) |
| South Africa |  | 0.86 |  | 24.66 |

**Supplementary Table S3.** Annual percentage change (95% CI) in truncated age-standardised colorectal cancer mortality by age at onset, population group and sex, 2002–2019 and 2011–2019. Group-specific rows exclude deaths with unrecorded population group. * p < 0.05.

| Onset | Group | Sex | 2002–2019 | 2011–2019 |
|---|---|---|---|---|
| Early (<50) | All groups | Both | +1.2* (0.5, 2.0) | +0.6 (−1.3, 2.6) |
|  |  | Male | +0.6 (−0.3, 1.5) | −0.4 (−2.4, 1.6) |
|  |  | Female | +1.9* (0.9, 2.9) | +1.7 (−0.5, 4.0) |
|  | Black African | Both | +2.9* (1.7, 4.0) | −0.2 (−2.6, 2.3) |
|  |  | Male | +1.5* (0.4, 2.7) | −1.4 (−3.8, 1.2) |
|  |  | Female | +4.2* (2.7, 5.8) | +1.0 (−1.8, 3.9) |
|  | White | Both | +2.9* (1.6, 4.2) | +3.9* (0.7, 7.1) |
|  |  | Male | +3.8* (1.7, 6.0) | +3.3 (−1.8, 8.6) |
|  |  | Female | +1.7 (−0.2, 3.6) | +4.4* (0.2, 8.7) |
|  | Coloured | Both | +3.8* (2.4, 5.2) | +6.0* (1.9, 10.3) |
|  |  | Male | +3.6* (2.1, 5.1) | +4.9* (0.0, 10.1) |
|  |  | Female | +3.7* (1.4, 6.2) | +6.7* (1.2, 12.4) |
|  | Indian/Asian | Both | +0.2 (−2.7, 3.2) | −1.2 (−9.7, 8.2) |
|  |  | Male | 0.0 (−2.8, 2.9) | −4.3 (−11.0, 2.8) |
|  |  | Female | +0.4 (−3.3, 4.4) | +3.2 (−10.0, 18.4) |
| Late (≥50) | All groups | Both | +0.9* (0.3, 1.4) | +1.1 (−0.2, 2.5) |
|  |  | Male | +1.4* (0.8, 2.1) | +0.9 (−0.5, 2.3) |
|  |  | Female | +0.5 (−0.2, 1.2) | +1.4 (−0.4, 3.2) |
|  | Black African | Both | +5.1* (4.2, 5.9) | +5.0* (2.8, 7.3) |
|  |  | Male | +5.5* (4.4, 6.6) | +4.6* (2.7, 6.5) |
|  |  | Female | +5.0* (3.9, 6.0) | +5.6* (2.3, 9.0) |
|  | White | Both | +0.1 (−0.5, 0.7) | −0.8 (−2.4, 0.8) |
|  |  | Male | +0.2 (−0.6, 1.0) | −1.3 (−3.6, 1.1) |
|  |  | Female | −0.1 (−0.7, 0.6) | −0.8 (−2.4, 0.8) |
|  | Coloured | Both | +2.9* (1.9, 4.0) | +2.1 (−0.2, 4.5) |
|  |  | Male | +3.8* (2.4, 5.2) | +2.6 (−0.4, 5.7) |
|  |  | Female | +2.3* (1.2, 3.4) | +1.9 (−0.7, 4.5) |
|  | Indian/Asian | Both | +1.5* (0.3, 2.6) | −1.5* (−2.7, −0.2) |
|  |  | Male | +1.8* (0.4, 3.3) | −1.5 (−3.8, 0.8) |
|  |  | Female | +1.1 (−0.1, 2.4) | −1.0 (−2.6, 0.7) |

